# Covid-19 Pandemic in relation to levels of Pollution with PM2.5 and Ambient Salinity – An Environmental Wake-up Call

**DOI:** 10.1101/2020.05.03.20087056

**Authors:** Yves Muscat Baron

## Abstract

**INTRODUCTION:** Covid-19 infection continues to be a source of great loss of life and global suffering, necessitating national lockdowns. There are however some countries and cities which appear to have contained the pandemic. Common to these countries are environmental factors including the levels of particulate matter (PM2.5) and ambient salinity.

**METHOD:** PM2.5 and ambient salinity were assessed in a number of cities, differentially affected by Covid-19 infection. The cities chosen to be assessed were divided into two groups. The first group included cities having significantly high rates of Covid-19 infection, while the second group consisted of coastal cities or small island countries, all of which have low rates of Covid-19 infection. Minimum and maximum levels of PM2.5 were obtained from the Air Quality Index, one month before and one month after statutory lockdown. Salinity levels were obtained from a study that assessed chloride ion wet deposition, a surrogate for ambient salinity.

**RESULTS:** One month prior to the statutory national lock-down or mandatory restrictions, there appear to be high levels of particulate matter, PM2.5 (min-max 67.4 - 118.7 AQI), in countries which had a high incidence of Covid-19 infection compared to lower levels in countries that have contained the infection (min-max 45.6 - 79.8) (p<0.046). One month after national restrictions there still appeared to be higher levels of particulate matter, PM2.5 (min-max 51 - 90.5 AQI), in countries which had a high incidence of Covid-19 infection compared to countries that have contained the infection (min-max 42.7 - 69.5 AQI) but this was not statistically significant.

There seemed to be an inverse relationship between Covid-19 infection and ambient salinity levels. Countries that were spared high Covid-19 infection rates, besides their geographical isolation, also have higher ambient salinity levels (124 - 617mgCl/m^2^/TAG) compared to salinity levels noted in countries with high Covid-19 rates of infection (28.4 - 162. mgCl/m^2^/TAG) (p<0.003).

**CONCLUSION:** High levels of PM2.5 in the presence of low ambient salinity may increase the risk of Covid-19 infection in the population. Addressing these two environmental factors may attenuate the severity of the pandemic.

## INTRODUCTION

Covid-19 infection continues to wreak havoc, in several populations with great loss of life, economies are at a standstill with unemployment increasing exponentially and millions are threatened with starvation. There are however ‘islands of hope’ such as the author's country, Malta, which appears to have contained the infection. There are other “islands of hope” such as the Hong Kong, Cyprus, South Korea, New Zealand and Taiwan.

There are common factors between these countries which appear to have contained the Covid-19 pandemic. The most obvious common factor is geographical, whereby most of these countries are in fact islands, except for South Korea which however, because of its geopolitical circumstances, can be considered an "island". This form of physical isolation allows the possibility of effective measures ensuring successful social distancing, preventing large population movements which may encourage spread of infection. National jurisdictions may be able to function more efficiently in small populations. Legislation enforcing public health measures such as social distancing and processes to protect the elderly and vulnerable individuals may be more effective at reaching the general citizenship of a nation^1^.

The epicentres of Covid-19 infection such as Wuhan, Qom and Bergamo have high population densities. Similarly in the USA, New York and New Orleans have both high population densities and high incidences of Covid-19 infection. This was initially thought to be a catalyst for the rapid Covid-19 spread; however there are other cities spared from Covid-19 with higher population densities^2^. Moreover, the countries mentioned earlier which appear to have contained the Covid-19 infection also have high population densities^3^. As a corollary to high population densities, pollution levels are relatively elevated in Wuhan, Qom and Bergamo as indicated by atmospheric particulate matter (PM2.5 and PM10), nitrogen dioxide and sulphur levels.

## METHODOLOGY

The particulate matter PM2.5 and ambient salinity were assessed from a number of cities, differentially affected by Covid-19 infection. The cities chosen to be assessed were divided into two groups. The first group (Group 1) included Wuhan, Qom, Bergamo, Paris, Madrid, London and New York, all having significantly high rates of Covid-19 infection. Group 2 consisted of cities including Seoul, Napoli, Brest, Eastbourne, Taipei, Hong Kong and small countries such as Malta and Cyprus, all of which have very low rates of Covid-19 infection (WHO Report 2020)^4^

Data on particulate matter (PM2.5) was obtained from the Air Quality Index, which is updated in real time giving minimum and maximum levels for each day of the month^5^. The levels of particulate matter (PM2.5) were calculated one month before and one month after national statutory lock-downs or mandatory restrictions were instituted. Eyeballing the colour coded depiction of the Air Quality Index the air pollution as regards particulate matter (PM2.5) was worse even two months before national statutory lock-downs or mandatory restrictions were instituted in Wuhan, Qom and Bergamo. However to retain a standard time-line of one month, the data one month before and after national statutory lock-downs or mandatory restrictions were assessed. Data from Qom before lockdown was not available so that of Teheran before and after lockdown was utilized.

Chloride Ion Wet deposition in mgCl/m^2^/Tag is used as an index of ambient salinity. Data for ambient salinity was obtained from an indexed colour coded map compiled by Fraunhofer ISE, through a site set up by the Galvanizers Association^6^. This site determines the level of salinity in different regions in the world in connection with the risk of automobile rusting. The data are given as a minimum and maximum level in mgCl/m^2^/Tag.

## RESULTS

There appears to be elevated levels of particulate matter, PM2.5 (67.4-118.7 AQI), in countries which had a high incidence of Covid-19 infection compared to lower levels in countries that have contained the infection (45.6-79.8) (p<0.046). One month after national statutory lock-down or mandatory restrictions there still appeared to be higher levels of particulate matter, PM2.5 (51-90.5 AQI), in countries which had a high incidence of Covid-19 infection compared to lower levels in countries that have contained the infection (42.7-69.5) but this was not statistically significant (p=n.s).

The countries that seem to have contained the pandemic are surrounded by sea, increasing ambient salinity. Countries that evaded high Covid-19 infection rates have higher ambient salinity rates 124-617mgCl/m^2^/TAG compared to 28.4-162.8 mgCl/m^2^/TAG (p<0.003) noted in countries sustaining high Covid-19 rates of infection.

The results are demonstrated in two tables designated Figure 1 and Figure 2.

**Fig 1.**
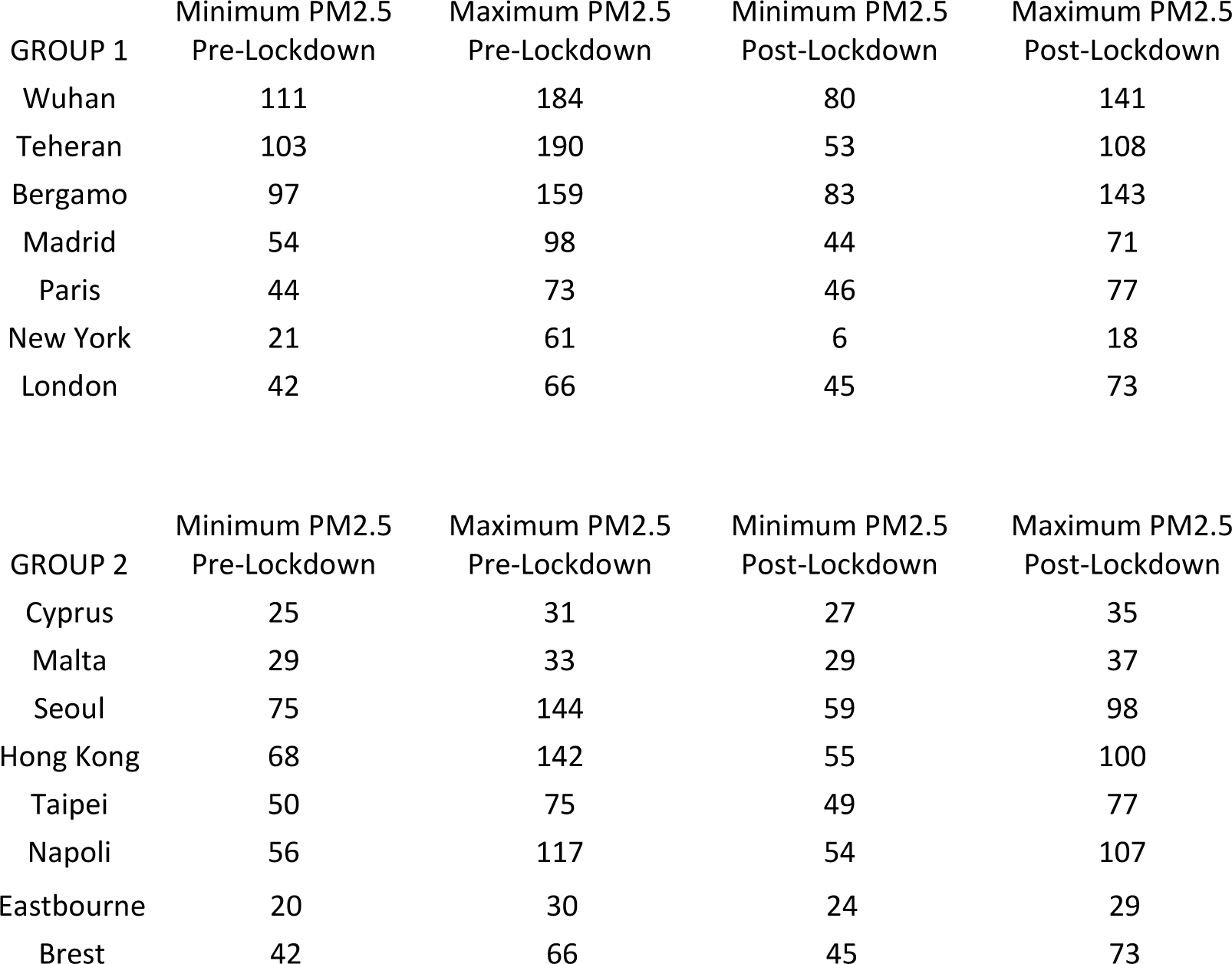
Particulate Matter 2.5 expressed in Air Quality Index (AQI), minimum and maximum levels, one month before and after lockdown. The p value between the two groups of countries/cities is p< 0.046. Group 1 had a higher incidence of Covid-19 infection and similarly had a higher PM2.5 AQI than Group 2. (Data from AQI Real time Air Quality Index^5^).

**Fig 2.**
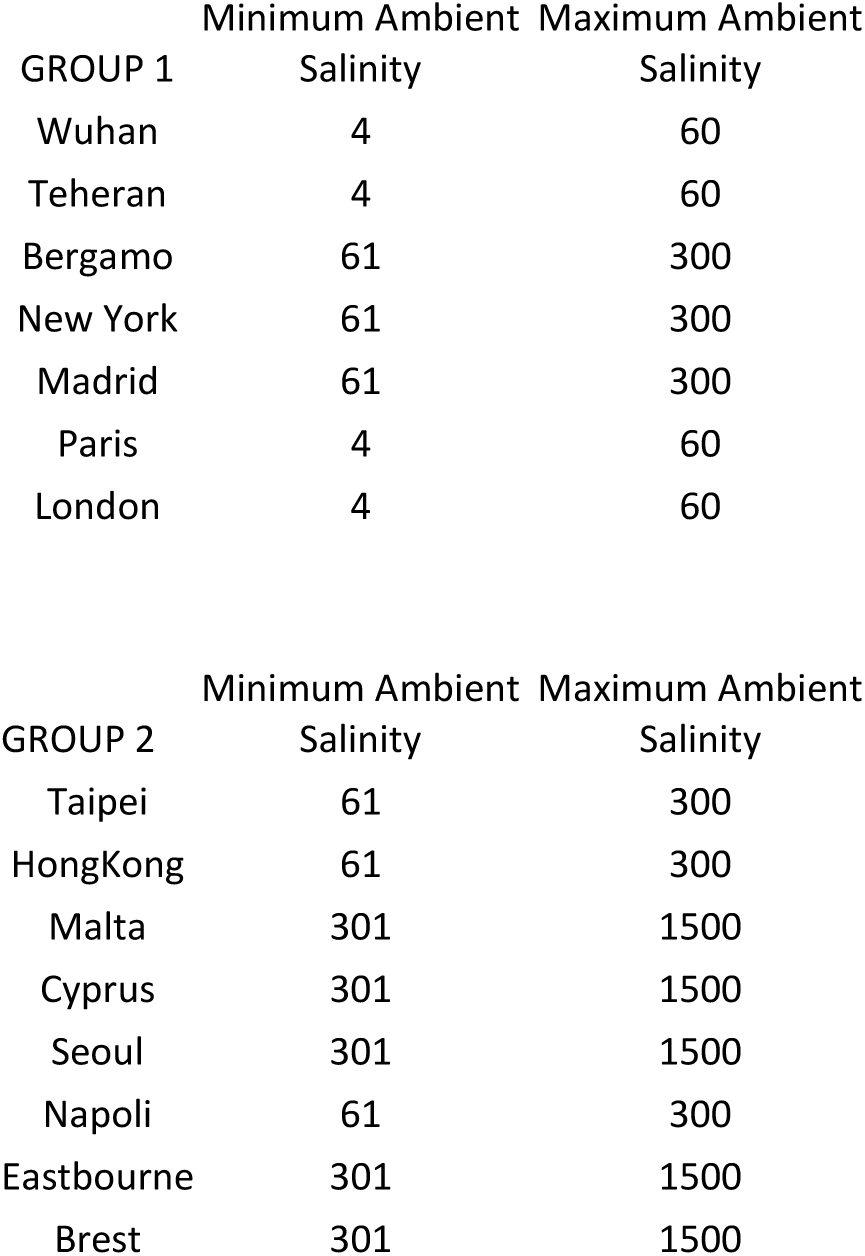
Ambient Salinity in mgCl/m^2^/Tag indicating minimum and maximum levels. The p value between the two groups of countries/cities is p< 0.003. Group 1 had a higher incidence of Covid-19 and similarly had a lower ambient salinity level in mgCl/m^2^/Tag compared to the Group 2. (Data indicating the Chloride Ion Wet Deposition, a variable reflecting the environmental salinity^6^).

## DISCUSSION

With the benefit of foresight, “the islands of hope” instituted social distancing. Besides reducing human to human viral transmission, physical distancing may have impacted pollution levels due to lower levels of economic activity. This was the case with Malta, whereby atmospheric pollution decreased by 70%. Before statutory lockdown, there may be synergism between high PM2.5 levels and Covid-19 infection. Addressing this synergism may protect the populations from airborne infections such as the Covid-19 infection and foster respiratory health. There appear to be strikingly elevated pollution levels of PM2.5 a few weeks prior to the outbreak of Covid-19 in Wuhan, Qom and Begamo^5^. These elevated levels of PM2.5 occurred during the winter months presumably due to combustion of fuel for heating purposes.

Another ambient factor may come into play in the form of the level of atmospheric salinity. A colour-coded map of the distribution of the earth's salinity has been provided by Fraunhofer-ISE indicating the Chloride Ion Wet Deposition, a variable reflecting the environmental salinity^6^. There may be a connection between the level of ambient salinity and the rate of Covid-19 infection.

The level of salinity as indicated by the colour-coded map confirms the elevated levels of Chloride Ion Wet Deposition in these countries. Wuhan has been noted to be one of the cities with the least salinity in the air^7^. Moreover, eyeballing the disease spread in some countries that have been hard hit by Covid-19 infection, there appears to be a correlation of Covid-19 infection rates and ambient salinity as in Qom and Bergamo. Other examples of this correlation are France and the U.K. The high salinity Brittany (Brest) region had fewer infection rates compared to the central (Paris) and eastern regions in France (lower salinity). Similarly, the South East region of the U.K. with higher saline levels had lower Covid-19 infection rates compared to London which appears to have a lower Chloride Ion Wet Deposition. This can also be seen in Italy whereby the saline deficient Lombardy region (Bergamo) was severely affected by Covid-19 infection, while the more saline rich south of Italy (Napoli) have very low infection rates.

Healthy levels of endo-tracheal sodium chloride protect the respiratory airways from disease. Ciliary action is improved when the saline content of the endobronchial tree is optimal. Bronchial mucous is less viscous with optimal levels of sodium chloride. In fact, patients with cystic fibrosis are administered hypertonic saline endobronchially to mobilize stagnant mucous plugs^8^.

The Covid-19 virus is hydrophobic and low concentrations of chlorinated solutions are lethal to the virus. Ambient salinity may be another environmental factor that may have protected some populations preventing airborne infection from the Covid-19^9^.

Besides the lethal effect of sodium chloride on the virus, there may be a connection with aerosol adhesion of sodium chloride with the pollutant PM2.5. In snowbound countries, where salt is applied to roads to encourage snow to melt, aerosolization of the salt together with PM2.5 has been noted^10^. The amalgamation of sodium chloride to PM2.5 may consequently attenuate the inflammatory effect of the polluting particle. Inflamed pulmonary tissue may act as an ideal pabulum for the Covid-19 virus to adhere to the epithelium and the presence of sodium chloride may inhibit viral adhesion and infective activity. Recent evidence suggests that Covid-19 may have “piggy-backed” on to PM2.5 particles to evade the bronchial tree defences and colonize the alveoli^11^. The presence of sodium chloride on PM2.5 particles may act as a deterrent against the concomitant adhesion of Covid-19 to these particles, protecting populations living in regions with high salinity.

From the above discussion, there may be a suggestion that the rapid and ruthless global advance of Covid-19 infection may have occurred due to the combination of a number of environmental factors besides the virulence of the virus. The increase in pollution in particular PM2.5, and lack of ambient salinity may have acted in concert to catalyse the pandemic. Measures to address these factors may prove useful to reverse the onslaught of the Covid-19 Pandemic. The statutory lockdown indicated that reduction in PM2.5 did occur when these restrictions were instituted. Reducing pollution, diminishing PM2.5 in particular, is imperative. Populations living in elevated levels of ambient salinity also appear to be protected. Instituting a synergistic approach of reducing pollution combined with “salinating” measures of habitable air, especially in enclosed spaces with nebulised saline, may protect populations from the further advancement of the pandemic.

## Data Availability

Data obtained from referenced publications and external data sets as shown below.

https://openknowledge.worldbank.org/handle/10986/5991

https://www.who.int/publications-detail/infection-prevention-and-control-during-health-care-when-novel-coronavirus-(ncov)-infection-is-suspected-20200125

http://www.ceic.data.com

http://www.medrxiv.org

